# Integrating Functional Protein Drug Target Data into a Precision Oncology Molecular Tumor Board

**DOI:** 10.1101/2024.10.17.24315608

**Authors:** Allison L. Hunt, Jamie Randall, Mahesh M. Mansukhani, Kara Nyberg, Aratara Nutcharoen, Justin Davis, Brian Corgiat, Claudius Mueller, Savannah Melvin, Meenakshi Sharma, Laura Johnston, Whitney Swain, Tamara Abulez, Nicholas W. Bateman, G. Larry Maxwell, John Deeken, Amin Benyounes, Emanuel F. Petricoin, Timothy L. Cannon, Thomas P. Conrads

**Affiliations:** Women’s Health Integrated Research Center, Women’s Service Line, Inova Health System, 3289 Woodburn Road, Annandale, VA 22003, USA; Gynecologic Cancer Center of Excellence and the Women’s Health Integrated Research Center, Department of Gynecologic Surgery and Obstetrics, Uniformed Services University and Walter Reed National Military Medical Center, 8901 Wisconsin Avenue, Bethesda, MD 20889, USA; Inova Schar Cancer Institute, Inova Health System, 8081 Innovation Park Drive, Fairfax, VA 22031, USA; Department of Pathology and Cell Biology, Columbia University Irving Medical Center, 630 W 168th Street, New York, NY 10032, USA; Department of Pathology, Inova Fairfax Hospital, 3300 Gallows Road, Falls Church, VA 22042, USA; Ignite Proteomics, Inc., 15000 W 6th Avenue, Golden, CO 80401, USA; The Henry M. Jackson Foundation for the Advancement of Military Medicine, Inc., 6720A Rockledge Drive, Suite 100, Bethesda, MD 20817, USA; Center for Applied Proteomics and Molecular Medicine, George Mason University, 10920 George Mason Circle, MSN 1A9, Manassas, VA 20110, USA

**Keywords:** proteomics, next generation sequencing, laser microdissection, cancer, reverse phase protein microarray, personalized medicine, molecular tumor board

## Abstract

Collaborative review of molecular profiling data by multidisciplinary molecular tumor boards (MTB) is increasingly important for improving patient management and outcomes, though currently relies nearly exclusively on nucleic acid next-generation sequencing (NGS) and limited panels of immunohistochemistry-based protein abundance data. We examined the feasibility of incorporating real-time laser microdissection (LMD) enrichment of tumor epithelium and commercial CLIA-based reverse phase protein array (RPPA) protein drug target expression/activation profiling into our cancer center’s MTB to complement standard clinical NGS-based profiling. The LMD-RPPA workflow was performed within a therapeutically permissive timeframe with a median dwell time of nine days, during which specimens were processed outside of standard clinical workflows. The RPPA-generated data supported additional and/or alternative therapeutic considerations for 54% of profiled patients following review by the MTB. These findings suggest that integrating proteomic/phosphoproteomic and NGS-based genomic data creates opportunities to further personalize clinical decision-making for precision oncology.

## Introduction

Cancer diagnostics and treatments have significantly improved in recent years due in part to the continuous development and optimization of highly sensitive technologies and molecular assays for improved early detection, routine screening, and therapeutic selection; collectively contributing to advancing personalized medicine approaches. Despite these advances, cancer remains the second leading cause of death in the United States with over two million new cancer diagnoses and 611,720 cancer-related deaths predicted to occur in 2024^1^. Molecular tumor boards (MTBs) are becoming more commonplace in clinical oncology, in which multidisciplinary committees comprised of clinical and research specialists review the clinical histories and molecular profiling data for each patient and identify the best therapeutic options available, evidenced by clinical trials or known tumor molecular signatures.

The molecular profiling data available from clinical trials is largely from next generation sequencing (NGS) studies correlating genomic and/or transcriptomic alterations with therapeutic responsiveness. Cancer therapeutics, however, largely exert their anti-cancer activity at the level of the proteome, either by targeting specific protein activities (e.g., trastuzumab targeting human epidermal growth factor receptor 2 (HER2) kinase signaling^2^), or by disrupting cellular processes mediated by proteins (e.g., gemcitabine inhibiting DNA synthesis and cell cycle progression^3^). Genomic variation and transcriptomic expression are at best loosely correlated with protein activity and/or abundance^4–7^, attributed partially to post-translational modifications (PTMs) that modify protein activity, the presence of multiple protein isoforms transcribed/translated from a single gene, and the post-translational stability and/or localization of a protein either within or outside of a cell. Currently, protein-based profiling in clinical oncology is essentially limited to diagnostic immunohistochemistry (IHC).

The translational (clinical) relevance of transcript-based signatures is often limited in part due to the fact that large-cohort studies predominantly identify signatures from bulk tissue samples containing variable admixtures of tumor, stroma, and immune cells present within the tumor microenvironment (TME)^8,9^, which complicates therapeutic selection. Recent provocative studies have demonstrated widespread biological “misinterpretation” of gene expression signatures due to cell admixture^10^, and include the identification of genes/proteins correlating with poor outcomes in cancer patients that are stromally-derived^5,11–15^. Tissue preparatory techniques that isolate tumor and/or non-tumor cell populations, such as laser microdissection (LMD) or single cell-based analyses (single cell sequencing or single cell proteomics), are needed to selectively and accurately identify cell-specific prognostic and/or theragnostic molecular profiles from the tumor tissue microenvironment (TME).

We recently demonstrated that of incorporating CLIA-based reverse phase protein array (RPPA) drug target mapping into a precision oncology MTB for therapeutic decision-making significantly increases both actionability frequency and patient outcomes^16,17^. Based on the promise of RPPA and/or other new multiplexed and quantitative proteomic platforms, which directly measure the expression and activation of actionable protein drug targets, we sought to build on this work by evaluating the operational feasibility of integrating our hyphenated LMD- RPPA CLIA workflow for a real-time multi-omic return of data in our cancer center’s precision oncology MTB.

## Results

### Clinical Characteristics of Cohort

A total of 174 patients with solid tumor malignancies seeking treatment were consented and enrolled into this study to examine the feasibility of incorporating a hyphenated workflow involving LMD enrichment of tumor epithelium for RPPA-based proteomic analysis (Figure 1, Supplemental Table 1). All patients were from an intention-to-treat population; therefore, the primary aim of this study was to determine whether the implementation of new multi-omic workflows into MTB discussions could be performed within a treatment-permissible timeframe, and to secondarily determine whether complementary RPPA-based data would reveal additional or alternative therapeutic options based on NGS and IHC data alone.

**Figure 1.**
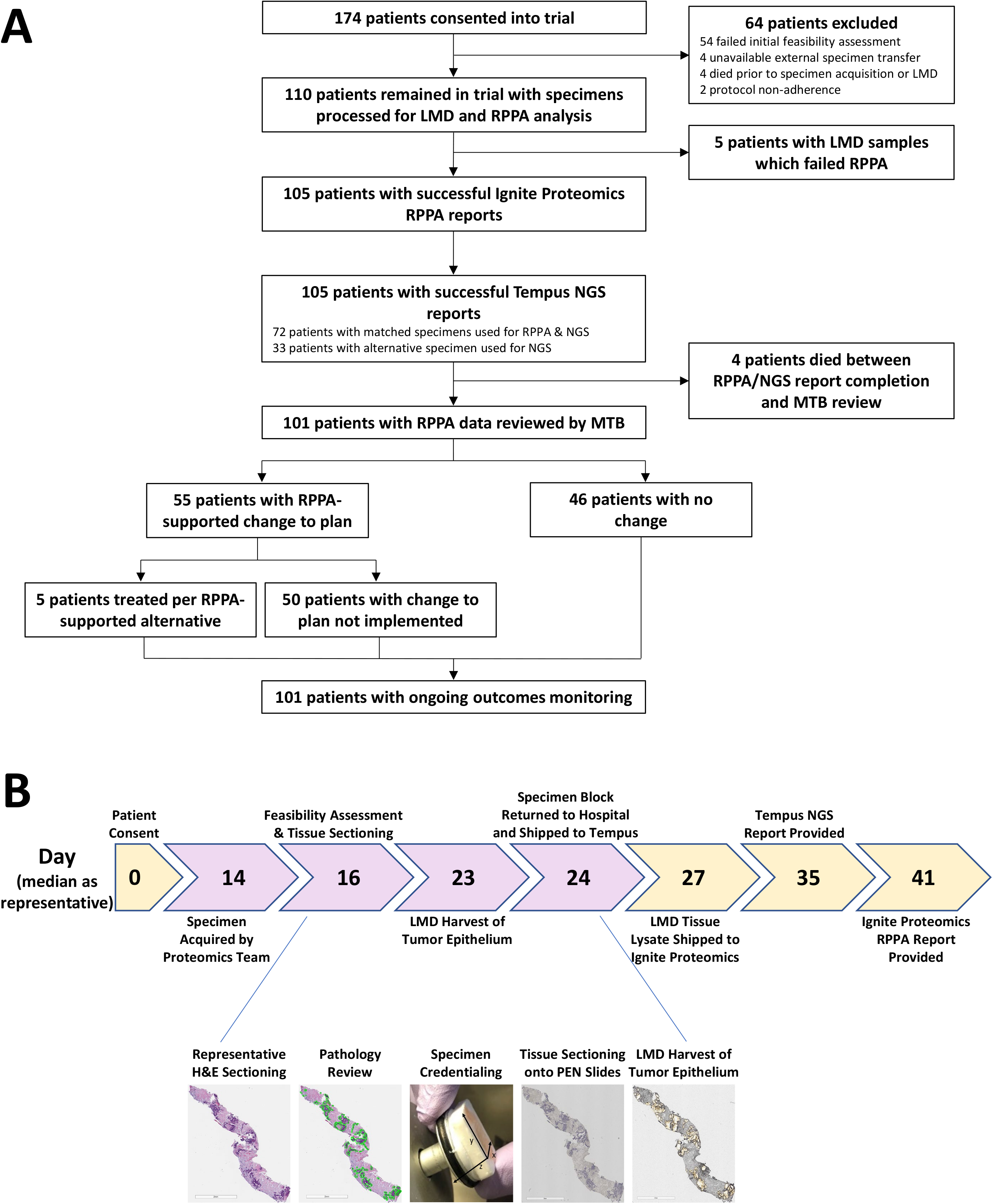
Workflow diagram. (A) Flowchart depicting the number of patients included or excluded at each timepoint of the study workflow. (B) Representative timeline and pre/post-LMD images. Purple arrows represent the days in which the specimen block was in the possession of the proteomic workflow team, i.e., outside the normal clinical MTB workflow.

Our cohort comprises 31 different primary malignancies, broadly categorized into 9 disease groups and 41 different histological cell types (Table 1). Most patients had gastrointestinal (n=77 patients, 44.3%), breast (n=26 patients, 14.9%), or skin (n=23 patients, 13.2%) tumors. Most tumors were adenocarcinomas (n=87 patients, 50%), though several tumors of rare histology were included, such as angiosarcomas (n=2), ganglioneuroma (n=1), inflammatory myofibroblastic tumor (IMT, n=1), leiomyosarcoma (n=1), Merkel cell carcinomas (n=2), melanocytic matrical carcinoma (n=1), pigmented dermatofibrosarcoma protuberans (“Bednar tumor”, n=1), and sebaceous carcinoma (n=1).

**Table 1.**
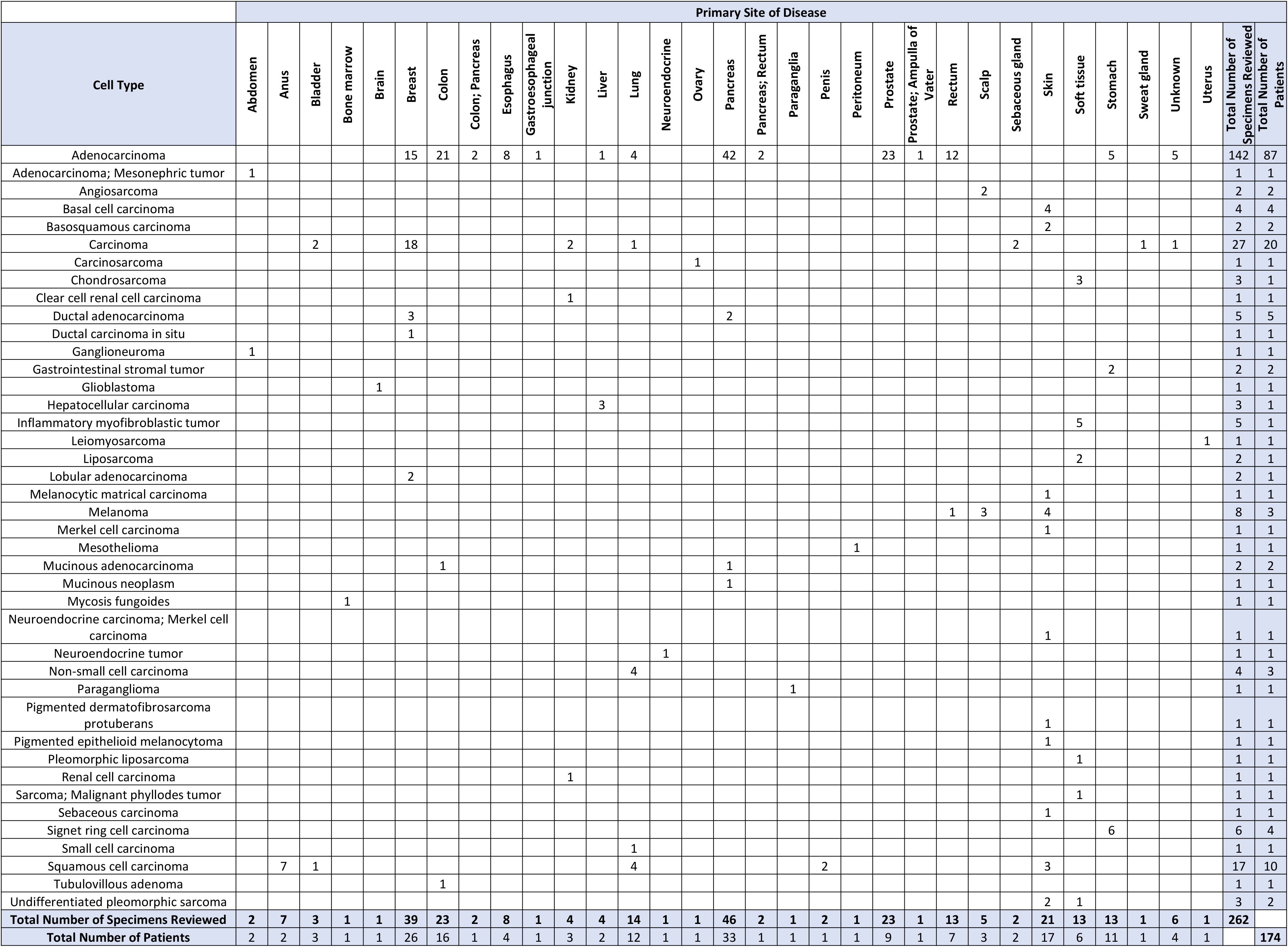
Total number of specimen blocks (n=262) from all patients (n=174) per cell type and primary site of disease.

### Patient Dropouts and Censors

During the feasibility assessment, patients were excluded if all available surgical specimens had insufficient tumor tissue to support LMD enrichment (n=54 patients) prior to RPPA analysis, if the patient died during the feasibility assessment period (n=4 patients), and/or prior to MTB review of the multi-omic data (n=4 patients), or if the relevant biopsy/surgery was performed at an external facility from which the tissue specimen or slides sectioned for LMD were unable to be obtained for analysis (“unavailable external specimen transfer”, n=4 patients) (Figure 1B, Table 2). Two patients (who were not lung cancer patients) were excluded when NGS was completed prior to RPPA analysis (“protocol non-adherence”). Further, despite achieving the target LMD harvest area (5-10 mm^2^ enriched tumor), five patients were excluded because the protein lysate concentration was insufficient for RPPA analysis. Three patients had multiple concurrent primary sites of disease, from each of which specimens were considered for analysis.

**Table 2.** Patient censors. Individual counts reported per primary site of disease and per censor classification. Percentage of patients censored per primary site of disease are reported in the bottom row and per censor reason in the right-most column.

|  | Primary Site of Disease |  |  |  |  |  |  |  |  |  |  |  |  |  |  |  |  |  |  |  |  |  |  |  |  |  |  |  |  |  |  |  |  |
| --- | --- | --- | --- | --- | --- | --- | --- | --- | --- | --- | --- | --- | --- | --- | --- | --- | --- | --- | --- | --- | --- | --- | --- | --- | --- | --- | --- | --- | --- | --- | --- | --- | --- |
| Patient Censored/Dropped | Abdomen | Anus | Bladder | Bone marrow | Brain | Breast | Colon | Colon; Pancreas | Esophagus | Gastroesophageal junction | Kidney | Liver | Lung | Neuroendocrine | Ovary | Pancreas | Pancreas; Rectum | Paraganglia | Penis | Peritoneum | Prostate | Prostate; Ampulla of Vater | Rectum | Scalp | Sebaceous gland | Skin | Soft tissue | Stomach | Sweat gland | Unknown | Uterus | Total Number of Patients | % of Patients |
| No | 1 | 1 | 2 | 1 | 1 | 18 | 10 |  | 4 | 1 | 2 | 1 | 3 | 1 | 1 | 15 | 1 |  |  |  | 5 | 1 | 6 | 1 | 2 | 11 | 4 | 4 | 1 | 2 | 1 | 101 | 58% |
| Yes (total) | 1 | 1 | 1 |  |  | 8 | 6 | 1 |  |  | 1 | 1 | 9 |  |  | 18 |  | 1 | 1 | 1 | 4 |  | 1 | 2 |  | 6 | 2 | 6 |  | 2 |  | 73 | 42% |
| Unavailable external specimen transfer |  |  |  |  |  |  | 1 |  |  |  |  |  |  |  |  | 1 |  |  |  |  |  |  |  | 1 |  | 1 |  |  |  |  |  | 4 | 2% |
| Patient death |  |  | 1 |  |  | 1 |  |  |  |  |  |  |  |  |  | 1 |  |  |  | 1 |  |  |  |  |  |  |  | 1 |  | 2 |  | 7 | 4% |
| Protocol |  |  |  |  |  | 2 |  |  |  |  |  |  |  |  |  |  |  |  |  |  |  |  |  |  |  |  |  |  |  |  |  | 2 | 1% |
| Specimen(s) inadequate | 1 | 1 |  |  |  | 5 | 5 | 1 |  |  | 1 | 1 | 9 |  |  | 15 |  | 1 | 1 |  | 4 |  | 1 | 1 |  | 5 | 2 | 5 |  |  |  | 59 | 34% |
| Specimen(s) inadequate; Patient death |  |  |  |  |  |  |  |  |  |  |  |  |  |  |  | 1 |  |  |  |  |  |  |  |  |  |  |  |  |  |  |  | 1 | 0.6% |
| Total Number of Patients | 2 | 2 | 3 | 1 | 1 | 26 | 16 | 1 | 4 | 1 | 3 | 2 | 12 | 1 | 1 | 33 | 1 | 1 | 1 | 1 | 9 | 1 | 7 | 3 | 2 | 17 | 6 | 10 | 1 | 4 | 1 | 174 |  |
| % Patients Censored/Dropped | 50% | 50% | 33% | 0% | 0% | 31% | 38% | 100% | 0% | 0% | 33% | 50% | 75% | 0% | 0% | 55% | 0% | 100% | 100% | 100% | 44% | 0% | 14% | 67% | 0% | 35% | 33% | 60% | 0% | 50% | 0% |  |  |

We sought to evaluate if there were associations between select tumor types and rates of patient dropout in our multi-omic workflow. Given the heterogeneous tumor origins observed in this pan-cancer study, only primary sites of disease with more than four patients were included in this assessment (Table 2). Among these, patients with primary tumors originating in the lung (9/12 patients, 75%), stomach (6/10 patients, 60%), and pancreas (18/33 patients, 55%) had the highest dropout rates. The lowest dropout rates were in patients with esophageal (0/4 patients, 0%), rectal (1/7 patients, 14%), or breast (8/26 patients, 31%) cancer. The most frequent reason for patient dropout was due to inadequate tissue availability for multi-omic analysis. No suitable tumor tissue specimens were available for NGS from two patients, so liquid biopsies (Tempus, Inc.) from these patients were profiled by NGS.

Alternative specimens for RPPA were selected when the amount of tissue available within a specimen block was insufficient (i.e. the necessary amount of tissue sections to achieve the target tumor LMD area could not be generated without exhausting the specimen). Among the inadequate tissue specimens, those obtained from lung (n=26 cassettes), lymph nodes (n=21 cassettes), liver (n=18 cassettes), pancreas (n=13 cassettes), and prostate (n=12 cassettes), were most commonly collected via fine needle aspirates (FNA) and had the highest rates of failure due to either minimal tissue depth within the specimen block or minimal tumor area for LMD enrichment (Supplemental Table 2). Of note, not all biopsies from these organs failed, as LMD enriched samples from lung, liver, pancreas, and prostate specimens were successfully obtained from other patients and analyzed in this study.

### Multi-omic Characterization of Tumors

Tumor epithelium was selectively harvested by LMD from 110 patients for quantitative proteomic analysis by RPPA, from which 105 successful RPPA and NGS reports were generated. The same tissue specimens were prioritized for multi-omic analysis by RPPA and NGS (using bulk tissue samples) whenever possible (Table 3). Matched specimens (i.e., same FFPE block) were used for 72/105 patients for RPPA and NGS analysis. Tissues from unmatched (different) specimen blocks were used for analysis by NGS vs RPPA from 33 patients. RPPA was used to quantify the abundance of 32 proteins and/or phosphoproteins with known cancer significance and/or that represent actionable drug targets^18,19^ (Supplemental Table 3). The RPPA and NGS reports from 101 patients were reviewed by the MTB. The multi-omic results from patients who died while the RPPA and/or NGS analyses were ongoing (4/105, 3.8%) were not reviewed by the MTB.

**Table 3.**
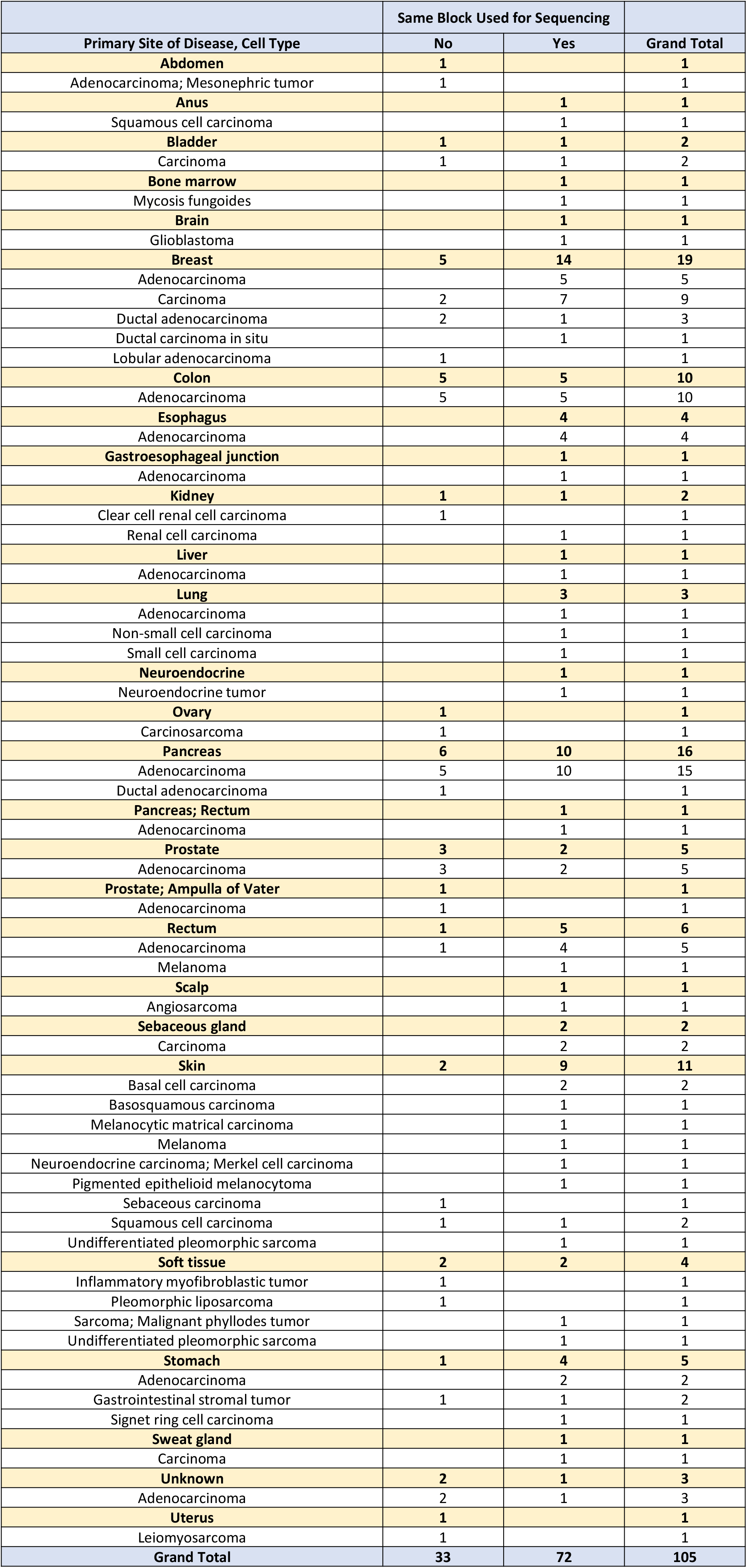
Matched specimens for RPPA and NGS. Matched specimens (“Yes”) for sequencing represent the same FFPE specimen block was used for both RPPA and NGS analysis. Counts of unmatched (“No”) specimens represent patients from whom different specimen blocks (and/or liquid biopsies, n=2 patients) were used.

Following specimen receipt by the proteomics team (median day 14 after patient consent; Supplemental Table 1), a representative H&E tissue section was reviewed and the tumor areas for LMD harvest were annotated and confirmed by a board-certified pathologist (median day 16 after patient consent; day 6 of the proteomic workflow). Specimens that passed this feasibility assessment (i.e. had sufficient areas of tumor to support LMD enrichment for RPPA and NGS) were further sectioned onto polyethylene naphthalate (PEN) membrane slides. The proteomic arm of the study required possession of each tissue specimen for a median of 9 days (day 24 after patient consent), during which time the specimen was not available to the hospital or NGS companies for standard clinical testing. Specimen blocks were returned to the hospital for NGS after generating tissue sections for LMD-RPPA or after failing the feasibility assessment. After tissue sectioning, the LMD-RPPA and NGS analyses occurred essentially in parallel (representative days 24-41 after patient consent), with the exception for patients with lung cancer or for specimens that failed the feasibility assessment for proteomic analysis and were therefore exclusively reserved for NGS analysis (Supplemental Table 1, Supplemental Table 4). NGS reports were available within a median of 11 days (representative day 35 after patient consent) after receipt of the FFPE tissue specimen by Tempus (Supplemental Table 4). RPPA reports were available within a median of 8 days (representative day 41 after patient consent) after receipt of the LMD sample lysate by Ignite Proteomics (Supplemental Table 1).

Given the heterogeneity of patients in this pan-cancer patient cohort, a preliminary analysis examining tumors associated with the highest and lowest protein recovery (µg/mm^2^ LMD tumor area) was performed. The median recovery across all samples was 0.574 µg/mm^2^ (Supplemental Table 5). Across tumor types, the highest protein recoveries were from tumors originating in the sebaceous gland (median=1.86 µg/mm^2^, n=2), neuroendocrine tumors (1.51 µg/mm^2^, n=1), and the gastroesophageal junction (1.21 µg/mm^2^, n=1), while the lowest recoveries were from tumors from the uterus (0.08 µg/mm^2^, n=1), lung (median=0.28 µg/mm^2^, n=4), and stomach (median=0.28 µg/mm^2^, n=6) (Supplemental Figure 1A). When categorizing the data by tissue sites sampled, biopsies obtained from the eyelid and conjunctiva (2.14 µg/mm^2^, n=1), omentum (median=1.33 µg/mm^2^, n=2), and colon (median=1.29 µg/mm^2^, n=2) had the highest recoveries, while those from the endometrium (0.08 µg/mm^2^, n=1), stomach (median=0.19 µg/mm^2^, n=3), and epiploic appendage (0.22 µg/mm^2^, n=1) had the lowest recoveries (Supplemental Figure 1B). Finally, when categorizing the data by tumor histology, neuroendocrine tumors (1.51 µg/mm^2^, n=1), gastrointestinal stromal tumors (GIST; median=1.34 µg/mm^2^, n=2), and pigmented epithelioid melanocytoma (1.28 µg/mm^2^, n=1) had the highest protein recovery, whereas leiomyosarcoma (0.08 µg/mm^2^, n=1), signet ring cell carcinoma (0.22 µg/mm^2^, n=1), and ductal adenocarcinoma (0.23 µg/mm^2^, n=1) had the lowest protein recoveries (Supplemental Figure 1C).

### Proteome-Informed Clinical Considerations

The multi-omic data from NGS and RPPA were reviewed by our cancer center’s MTB for 101 patients to generate a personalized list of considerations for later-line therapy based on currently available targeted therapies and clinical trials. The expression and/or activation levels for each of the 32 RPPA analytes were reported as a continuous percentage ranging from non-expressed/non-activated (0%) to highly expressed/highly activated (100%), and as a normalized intensity integer representing normalized percentage quartiles as follows: 0 (non-expressed/non-activated), 1+ (low expression/activation), 2+ (moderately expressed/activated), or 3+ (highly expressed/activated) (Figure 2, Supplemental Table 6), as previously described^19,20^. Broadly, highly abundant (2+ or 3+) RPPA targets and/or select less abundant (1+) RPPA targets predictive of a select therapeutic based on clinical trial outcomes were prioritized for a patient’s treatment regimen.

**Figure 2.**
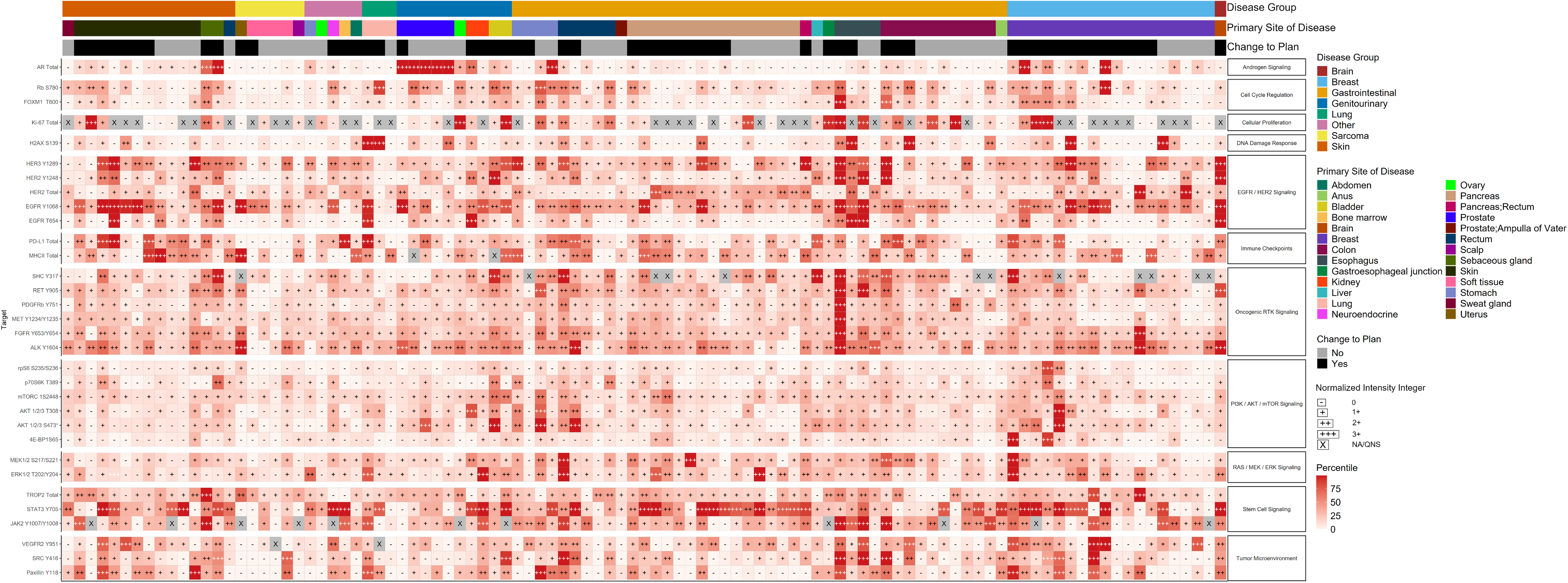
Additional and/or alternative therapeutic considerations (“change to plan”) based on RPPA-derived quantitative proteomic and/or phosphoproteomic data. Heatmap depicting the RPPA-quantified expression and/or activation of 32 protein and phosphoprotein analytes with known relevance as druggable targets in solid tumors relating to androgen signaling, cell cycle regulation, cellular proliferation, DNA damage response, EGFR/HER2 signaling, immune checkpoints, oncogenic RTK signaling, PI3K/AKT/mTOR signaling, RAS/MEK/ERK signaling, stem cell signaling, and the tumor microenvironment. NA/QNS = not applicable or quantity not sufficient, for RPPA abundances that were not reported for a given patient.

Across all disease sites, the RPPA-derived data supported additional or alternative therapeutic modalities for 55/101 patients (54.5%) (Figure 2, Supplemental Table 6). Prioritizing groups with ≥4 successfully reviewed patients, the primary sites of disease with the highest rates of recommended changes to therapy included breast (13/18 patients, 72.2%), skin (7/11 patients, 63.6%), and pancreas (9/15 patients, 60%). Patients with prostate (1/5 patients, 20%), soft tissue (1/4 patients, 25%), and rectal (2/6 patients, 33.3%) tumors were least likely to have alternative therapies recommended based on the RPPA workflow and/or the specific antibody panel used for analysis.

At the time of publication, the recommended changes had not yet been implemented because most patients had not exhausted their standard of care therapeutic options. Patient outcomes that result with/without implementation of the RPPA-informed changes will be evaluated and presented in the future.

### Recommendations for Integrating Proteomic Workflows into Clinical Practice

The most common limitations encountered while piloting this integrated multi-omic clinical MTB workflow are described in Table 4. Miniscule tissue volumes within specimen blocks and/or low tumor cellularity of the biopsied tissue (i.e., “inadequate tissue specimen(s)”) most frequently prevented proteomic/multi-omic analysis. The amount of tissue needed for each workflow (RPPA, DNA-seq, and/or RNA-seq) and a contextual representation of what that physically equates to in clinical practice per disease and biopsy tissue source should be carefully communicated between the research and surgical/interventional radiology teams prior to adoption of proteomic workflows into MTBs and/or prior to surgery/biopsy. If the pre-biopsy tumor size permits and if clinically safe for the patient, additional tissue collections to obtain multiple biopsy cores as well as either embedding all cores into a single FFPE specimen block or pooling of tissue from individual cores embedded into multiple blocks may help reduce the number of specimens that dropout due to inadequate tissue availability.

**Table 4.** Recommendations, limitations, tips and tricks, and lessons learned from pilot integration of proteomic workflows into the molecular tumor board.

| Limitation | Recommendation |
| --- | --- |
| Inadequate tumor area / tissue availability | Obtain additional biopsy cores at time of initial surgery and combine the microdissected areas from multiple cores and/or related specimen blocks for analysis. |
|  | Pre-screen biopsy specimens to select specimen block with most tissue and/or highest tumor content. |
| | Minimize the amount of tissue discarded while “facing” the specimen block prior to thin sectioning (i.e., “face” the block with the microtome blade thickness set to a maximum of 2-3 μm). Minimize the amount of tissue lost while generating ribbons. |
| Dropouts for “unavailable external specimen transfer” | Establish SOP’s / MTA’s with neighboring clinical facilities which may possess surgical specimens to ensure that blocks from outside facilities can be obtained (or that they are able to perform the tissue sectioning at their facility). |
| **General Tips & Tricks, Lessons Learned** |  |
| Collect LMD tissue into PCT tubes, then place the capped PCT tube into a 0.5 ml PCR tube for sample lysis to minimize evaporation. Transfer the sample lysate to o-ring screw top tubes prior to shipment on dry ice. |  |
| RPPA analyte panel selection: can prioritize general cancer-related targets or disease-specific panels. |  |
| Generate sections for LMD-RPPA before NGS for efficiency / availability of tissue after consumption for workflow. Consider commercial sequencing companies that require fewer numbers of sections and have higher sensitivity assays. |  |
| Annotations of tumors areas for LMD enrichment should be as precise as possible, minimizing white space and infiltrating stroma/lymphocyte areas, to most accurately predict the number of sections needed for LMD. Circling of large general tumor regions (as is common practice for many pathologists) is not recommended. After precise annotation, further scale-down the estimated obtainable area by a conservative ratio (ex. assume 60-80% of the tumor area is retained in serial sections and can be obtained) to account for tissue variability between sections (exhaustion of tumor areas) or other cutting artifacts that may occur (ex. tissue curling/tearing). |  |

Care should be taken during histopathology sectioning when generating tissue sections, particularly for specimens with minimal starting material, such as thin needle biopsies. Specifically, while “facing” the tissue specimen (i.e., shaving off the top layers of the FFPE block to obtain a full tissue section representing the complete intact cross-sectional area), histopathology technicians should set the microtome blade to a minimum thickness to minimize the loss of tissue.

## Discussion

Genomic variation and transcriptomic expression are imperfect and indirect proxies of protein drug target expression and/or activation. Accurate and sensitive quantitative and multiplexed measurements of proteins and phosphoproteins are increasingly being acknowledged as a missing component of molecularly informed treatment planning. Moreover, recent studies have shown that pre-treatment levels of tumor proteins and phosphoproteins are highly predictive of response in the absence of and independent of underpinning genomic alterations^20–30^. A critical need thus exists to identify and validate proteomic and/or phosphoproteomic biomarkers with diagnostic, prognostic, predictive, and/or theragnostic importance, which may only occur through the incorporation of proteome-focused analyses into clinical practice. We therefore present our experience incorporating real-time proteomic testing via RPPA analysis into our cancer center’s MTB workflow, for which we demonstrate alignment of proteomic/multi-omic analyses within a treatment-permissible timeframe and the potential for significant clinical benefit. Specifically, collaborative review of the proteomic data in MTB discussions resulted in identification of additional and/or alternative treatment considerations for 54% of the patients who had been successfully profiled and only prolonged standard clinical workflows by a median of nine days.

Integrating our hyphenated LMD-RPPA workflow into our cancer center’s precision medicine approach facilitated several anecdotal discoveries with profound clinical significance. One patient was diagnosed by IHC and standard clinical diagnostics with an anaplastic lymphoma kinase (ALK)-positive inflammatory myofibroblastic tumor^19^. RPPA-derived proteomic/phosphoproteomic data on the LMD enriched tumor revealed that ALK signaling was not activated (i.e., not phosphorylated) in the patient’s tumor after treatment with an ALK-directed tyrosine kinase inhibitor (TKI), thereby contraindicating the continued use of subsequent generations of ALK TKI. This indication was further supported by the presence of a secondary *ALK* mutation that was correlated with reduced ALK TKI sensitivity *in vitro*.

Another patient on this study had recurrent triple negative breast cancer (TNBC) and progressed on five prior lines of therapy prior to RPPA analysis (manuscript accepted, pending publication). Contrary to the IHC-derived evidence as being HER2-negative, the RPPA data from the LMD enriched tumor specimen revealed substantial HER2^Total^ and phosphorylated (p)HER2^Y1248^ and pHER3^Y1289^ levels, which supported the use of a HER2-directed therapy. The patient had a complete response to 6^th^ line treatment with trastuzumab deruxtecan (T-DXd), a therapy that would have not been otherwise selected based on standard clinical diagnostics alone. The LMD enriched tumor samples from several patients with pancreatic adenocarcinoma (PDAC) enrolled into this study were found to have elevated HER2^Total^ expression, comparable to the levels observed in breast cancer patients with HER2-positive and HER2-low tumors^31^.

While not significantly activated (phosphorylated), the high rate of elevated HER2^Total^ expression in PDAC tumors provided foundational evidence supporting further studies examining the use of HER2-targeting antibody drug conjugates (ADCs) in this patient population, for whom existing therapeutic options are limited and clinical outcomes are dismal. Further, observations of protein expression and/or activation patterns that diverge from current or dogmatic understanding, such as this finding, may support a need for selective RPPA panels that align with known disease biology per cancer type into MTB proteomic/multi-omic workflows.

While the survival outcomes for cancer patients continue to gradually improve overall, current estimates predict only a 69% 5-year survival rate for all cancers, and the outcomes for patients with pancreatic, liver, esophageal, and lung cancers remain far poorer^32^. There is no specific efficacy threshold required for FDA approval of new drugs or new drug indications. The median response rate endpoint for approval for recent drugs and/or indications is ∼41% with a median response duration of only 9 months ^33^. These low efficacy rates critically underscore the necessity for better therapeutic options. Our data support the findings of recent studies such as the I-SPY 2 trial^21,22^ to consider treating malignancies as “molecular target-oma’s” based on multi-omic profiling data (to specifically include proteome/phosphoproteome-level characterization of enriched tumor cell populations) in a personalized medicine manner, rather than strictly on the basis of disease origin site and histology. Specifically, the interpatient heterogeneity observed supports the need for treating each patient as an “n-of-1” using individualized molecular profiling for precision therapeutic decision-making. We successfully demonstrate that incorporating real-time LMD enrichment of tissue specimens and subsequent RPPA analysis as an orthogonal dataset to complement NGS data from bulk tissue specimens can be performed within a clinically-permissive timeframe and often yields additional and/or alternative therapeutic options when the multi-omic data are collectively reviewed in the MTB setting.

## Methods

### Patient Cohort

From October 2021 to December 2023, a total of 174 adult (≥18 years of age) patients with advanced solid tumor malignancies seeking treatment were consented and enrolled under a Western IRB-approved protocol into the Inova Schar Cancer Institute’s Molecular Tumor Board (MTB) study (protocol number U20-19-4308). The written informed consents included the provision to analyze and publish information and data regarding the patients’ clinical presentation, and results and data from precision medicine/next generation genomic sequencing testing, including the RPPA profiling data. Eligible patients had pathology-confirmed cancer by tumor biopsy and had tumor available for testing. Patients with multiple concurrent malignancies were eligible. Patients had received varied lines of previous therapy, ranging from treatment-naive to heavily pre-treated. All experimental protocols involving human data in this study were in accordance with the Declaration of Helsinki and written informed consent was obtained from all patients.

A total of 262 formalin-fixed, paraffin-embedded (FFPE) surgically-resected and/or biopsied tissue specimens were obtained from patients (average 2 blocks/patient, range 0-10). The specimens were acquired through surgical resections, excisions, computed tomography (CT)-guided biopsies, fine needle aspirations (FNA), punch biopsies, and/or bone biopsies. Liquid biopsies were used exclusively for NGS if all available solid tissue specimens were of insufficient quality and/or quantity.

Associated costs per specimen for in-house histology and LMD tissue processing as well as RPPA analysis were covered by the study at no cost to the patient. Costs associated with NGS were billed to the patient as a part of their standard of care treatment, with costs varying based on the specific assays ordered, insurance coverage, and financial assistance.

### Feasibility Assessment

After initial clinical diagnostic testing, the tissue specimens were assessed to determine the total amount of available tissue within a specimen block (width and depth of tissue) and the amount of specific tumor area available for enrichment via LMD. This determination was used to prioritize specimens for real-time quantitative analysis via a commercially available CLIA RPPA-based assay that measured 32 protein/phosphoprotein drug targets^18,19,31^ followed by real-time NGS for DNA-seq and/or RNA-seq.

To assess feasibility, a representative hematoxylin and eosin (H&E)-stained thin section for each case was scanned using the Aperio ScanScope XT slide scanner (Leica Microsystems). Foci of target tumor epithelium on the representative H&E slide were precisely annotated in Aperio eSlide Manager and Aperio ImageScope (Leica Microsystems) and confirmed by a board-certified pathologist. Specimens that contained sufficient tumor area and tissue depth within the FFPE block were credentialed for downstream workflows. Subsequent serial sectioning was performed with the highest priority of allocating tissue for NGS then RPPA. Chronologically (based on pre-defined optimal workflow outlined in the study protocol) tissue sections were first generated and allocated for the quantitative proteomic and phosphoproteomic workflows (RPPA), after which the specimen blocks were shipped to Tempus Labs, Inc. (Chicago, IL, USA) for NGS. Per institution preference, specimens from patients with lung cancer went directly from biopsy to NGS (prior to proteomic feasibility assessment) to accelerate treatment-based decision-making for administering EGFR inhibitors as indicated. Specimens that failed the feasibility assessment for LMD enrichment prior to RPPA-based proteomic analysis (i.e., had insufficient target tumor area per section and/or tissue thickness) but had sufficient tissue for NGS alone were shipped to Tempus and an alternative specimen block was requested, if available, for additional review for inclusion in the RPPA workflow. Specimens that failed the feasibility assessment for both the proteomic and/or NGS workflows were returned immediately, without testing being performed.

### Tissue Specimen Preparation for Laser Microdissection

A total of 111 FFPE tissue specimens were sectioned (8 µm) onto PEN membrane slides for LMD enrichment. The number of total slides was calculated from the number of sections needed to obtain a total cross-sectional area of enriched tumor epithelium of 5-10 mm^2^ for RPPA (median 2 slides/block, range 1-11, n=111 specimens). All slides were stained prior to LMD enrichment as previously described^5^. Briefly, slide preparation for LMD prior to RPPA did not involve counter-staining with eosin after hematoxylin but included color development in Scott’s Tap Water (Thermo Fisher Scientific, Inc.) and two final drying steps in xylenes. One representative LMD tissue section per specimen was imaged before and after LMD for each workflow for quality assurance and quality control (QA/QC) purposes.

### Laser Microdissection

LMD enrichment of tumor epithelium was performed on the LMD7 (Leica Microsystems). LMD tumor samples for RPPA analysis were collected in real-time (as patients were consented into the study and the specimens passed the feasibility assessment) into cylindrical pressure cycling technology (PCT, Pressure Biosciences, Inc.) microtubes. A lysis/extraction buffer was added at a ratio of 2.5 µl buffer per mm^2^ LMD tissue, as previously described^31^. The PCT microtubes were capped with PCT microcaps and placed into 0.5 ml PCR tubes, briefly centrifuged, and stored at -80 until sample lysis.

### Reverse Phase Protein Microarray

LMD-enriched tumor samples destined for RPPA analysis were heated at 95 for 20 min, briefly centrifuged, and heated at 80 for 2 h. After heating, the tubes were stored at 4 for one min and then centrifuged for 14,000 rpm for 15 min. The lysate supernatants were transferred to fresh low protein binding tubes and stored at -80 until shipment on dry ice to Ignite Proteomics (Golden, CO, USA) for RPPA analysis.

A 32-marker, CLIA-based RPPA analysis was performed as previously described^34^ to examine expression and activation (phosphorylation) of well-known actionable protein drug targets with known clinical relevance in solid tumors^18,19,31^. Briefly, sample lysates were heated and robotically arrayed onto nitrocellulose-backed slides that were probed with primary antibodies and a biotinyl tyramide amplification system to generate a fluorescent signal proportional to the analyte abundance. Signals were normalized to total protein and compared to a reference population to generate percentile and quartile scores in a patient report format. A minimum sample concentration of 100 µg/ml was required for quantification by RPPA. Multiple depositions were made to print samples with concentrations below this threshold, when possible.

### Next Generation Sequencing

Commercially available NGS was performed by Tempus Labs, Inc. DNA-seq was performed using the Tempus xT 648 gene panel^35^. Additional NGS testing (i.e., RNA-seq) was performed per physician discretion (Tempus xF 105 gene panel for cell-free DNA (cfDNA)^36^, PD-L1, tumor origin, etc.) or diagnosis. The samples for DNA-seq were prepared from tissue sections with an estimated 52% median tumor cellularity (range = 20-90%) after macrodissection/microdissection.

### Molecular Tumor Board Review

The NGS profiling data for each patient, along with demographic metadata including disease, age, and sex, was initially reviewed to evaluate therapeutic treatment considerations at our cancer center’s weekly Molecular Tumor Board (MTB) meeting attended by medical oncologists, molecular pathologists, and clinical trials research staff. After initial NGS review, the RPPA-based proteomics and NGS results were reviewed together with the relevant clinical histories to arrive at therapeutic considerations for each patient by the MTB. The MTB discussions were held biweekly and were attended in a hybrid virtual and in-person format by specialists from the clinical and research teams. The clinical team consisted of board-certified medical oncologists and pathologists, medical oncology fellows, genetic counselors, and clinical trials research staff. The research team consisted of subject-matter expert scientists specializing in molecular profiling (proteomics, phosphoproteomics, genomics, and/or transcriptomics), as well as histopathology and research technicians. A minimum of two board-certified medical oncologists were required to discuss the clinical results, interpret the multi-omic profiling data, and provide an MTB-informed consensus treatment recommendation based on the molecular profiling results. The clinical history and molecular profiling reports for each patient were first presented individually to the MTB in a lecture-style format led by a board-certified medical oncologist, after which the discussion was opened to a roundtable format to facilitate input and open dialogue between members of the clinical and research teams. After discussion and interpretation of the multi-omic data, an MTB-informed consensus treatment recommendation was recorded, provided to the treating physician, included in patients’ electronic medical record, and it was documented if there was a change from the original MTB NGS-informed treatment recommendations.

### Bioinformatic and Statistical Analysis

Descriptive statistics were used to summarize the clinical characteristics of the patients. Turnaround time metrics were calculated using 179 specimens obtained from 119 unique patients, unless otherwise specified. Specimens were excluded from all or some of the turnaround time calculations for one or more of the following reasons, as appropriate: (1) the patient was pre-defined as a “lead-in case” to facilitate early workflow transfers, (2) the specimen acquisition and/or analytical workflows for the patient did not adhere to the intended chronological workflow schema (e.g., NGS was performed prior to RPPA), (3) additional specimens were requested for clinical and/or research-related reasons (e.g., to examine pre- vs. post-treatment changes), (4) the related workflow step was hindered by reliance on a single vacant staffing position, or (5) the RPPA analysis was impacted by a universal commercial shortage of one of the required reagents and could not be performed until the supply issue was resolved. Box-and-whisker plots were generated using BoxPlotR^37^. The RPPA abundance/activation level heatmap was generated in R (version 4.4.0) with ggplot2 (version 3.5.1) and patchwork (version 1.2.0).

### Data Availability

The NGS data that support the findings of this study were generated by Tempus Labs, Inc. but restrictions apply to the availability of these data, which were used under license for the current study, and so are not publicly available. Data are available from the authors upon reasonable request and with the permission of Tempus Labs, Inc.

## Acknowledgements

This work was supported, in part, by a generous contribution from the Win Sheridan family funds. The authors would like to acknowledge Dr Paulette Mhawech-Fauceglia for histopathology image analysis support and Dr Kelly Conrads for critically reviewing the manuscript. Disclaimer: The views expressed in this article are those of the author(s) and do not necessarily reflect the official policy or position of the Uniformed Services University of the Health Sciences (USUHS), Department of the Navy, Department of the Air Force, Department of the Army, Department of Defense, or the United States Government. Mention of trade names, commercial products, or organizations does not imply endorsement by the U.S. Government.

## Author Contributions

Study concept and design: T.L.C., T.P.C., J.R., E.F.P. Data acquisition, analysis, and interpretation: A.L.H., M.M.M., K.N., J.R., A.N., J.D., B.C., C.M., S.M., M.S., L.J., W.S., T.A., N.W.B., G.L.M., J.D., A.B., E.F.P., T.P.C., T.L.C. Writing (original draft): A.L.H. Writing (review and editing): A.L.H., J.R., E.F.P., T.P.C., T.L.C. All authors gave final approval of the completed work and are accountable for accuracy and integrity.

---

Supplemental Table 1. Cohort metadata. Specimen-level data metrics for every tissue block received and reviewed as part of the proteomic workflow.

Supplemental Table 2. Inadequate specimen dropouts by primary site of disease and organ site sampled.

Supplemental Table 3. RPPA analysis antibody information. Rb (host) = rabbit. Ms = mouse. mAB = monoclonal antibody.

Supplemental Table 4. Turnaround time metrics including time from the date of patient consent, date of specimen acquisition by Tempus for NGS analysis, and date of NGS report availability. Summaries are provided only for patients who had available Ignite RPPA reports from either matched or unmatched specimens, and who were included in the proteomic workflow turnaround time calculations.

Supplemental Table 5. Protein recovery (µg/mm^2^) for all LMD tumor samples.

Supplemental Table 6. RPPA-derived proteomic and phosphoproteomic abundances and MTB- informed consensus decisions of additional and/or alternative therapeutic recommendations. RPPA abundances are reported as normalized intensity quadrant integers (0, 1+, 2+, 3+) and as percentiles (0-100%) relative to the reference population.

**Supplemental Figure 1.**
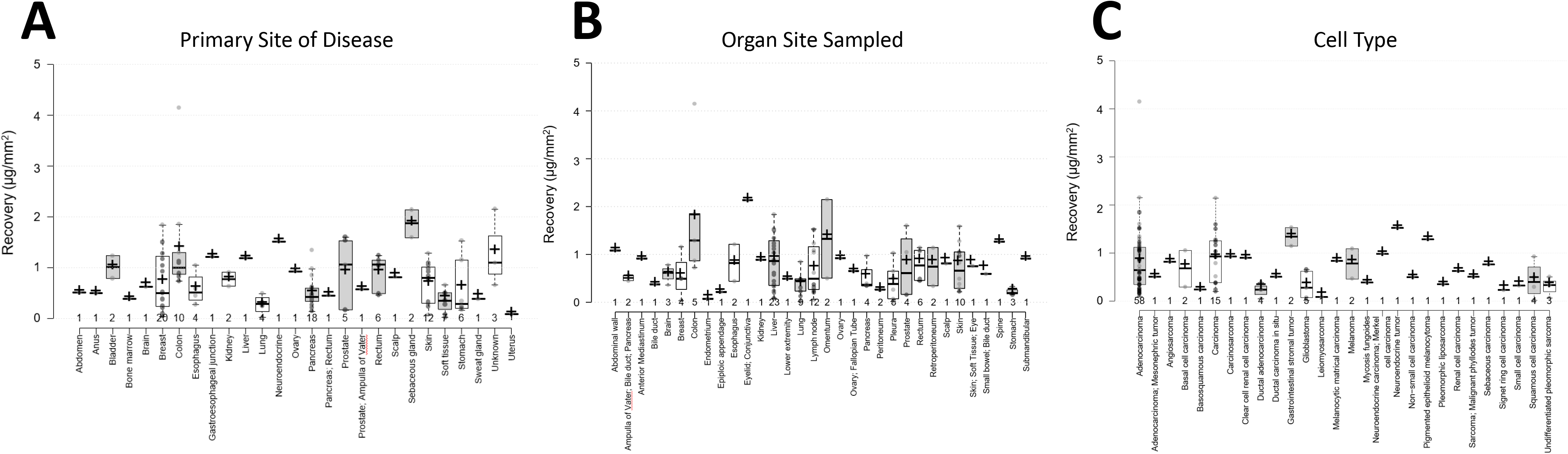
Box-and-whisker plots depicting protein recovery (µg/mm^2^) by (A) primary site of disease, (B) organ site sampled, and (C) cell type.

